# IE-Map: A novel in-vivo atlas and template of the human inner ear

**DOI:** 10.1101/19011908

**Authors:** Seyed-Ahmad Ahmadi, Theresa Marie Raiser, Ria Maxine Rühl, Virginia Lee Flanagin, Peter zu Eulenburg

**Author notes:** these authors contributed equally to this work (Seyed-Ahmad Ahmadi, Theresa Marie Raiser).

## Abstract

Brain atlases and templates are core tools in scientific research with increasing importance also in clinical applications. Advances in neuroimaging now allowed us to expand the atlas domain to the vestibular and auditory organ, the inner ear. In this study, we present IE-Map, an in-vivo template and atlas of the human labyrinth derived from multi-modal high-resolution magnetic resonance imaging (MRI) data, in a fully non-invasive manner without any contrast agent or radiation. We reconstructed a common template from 126 inner ears (63 normal subjects) and annotated it with 94 established landmarks and semi-automatic segmentations of all relevant macroscopic vestibular and auditory substructures. We validated the atlas by comparing MRI templates to a novel CT/micro-CT atlas, which we reconstructed from 21 publicly available post-mortem images of the bony labyrinth. Templates in MRI and micro-CT have a high overlap, and several key anatomical measures of the bony labyrinth in IE-Map are in line with micro-CT literature of the inner ear. A quantitative substructural analysis based on the new template, revealed a correlation of labyrinth parameters with total intracranial volume. No effects of gender or laterality were found. We provide the validated templates, atlas segmentations, surface meshes and landmark annotations as open-access material, to provide neuroscience researchers and clinicians in neurology, neurosurgery, and otorhinolaryngology with a widely applicable tool for computational neuro-otology.

## Introduction

The pervasiveness of brain imaging techniques across many disciplines has increased the relevance of standardized anatomical templates, including atlases of morphological and functional substructures. Almost the entire human central nervous system has been mapped: various templates and atlases based on MRI have charted out the human cortex, the subcortical nuclei, the thalamus and the cerebellum^1–4^, and more recently the brain stem and spinal chord^5^. These templates have found wide utility in group comparisons and quantitative assessments of individuals, becoming fundamental tools in neurology^6,7^, in neurosurgery^8,9^ and in the neurosciences^10^. Compared to whole-brain imaging, however, few studies have proposed templates and corresponding atlases to chart the anatomy of sensory organs, like the inner ear. Those that do exist have limited utility for non-invasive in-vivo comparisons, as they are based on imaging techniques that require radiation or ex-vivo preparation of tissue specimens, such as (micro-)CT^11^.

The inner ear contains several important substructures with specific functions^12,13^. The three semicircular canals detect head rotation, along with their ampullae that shelter the cupula containing the sensory hair cells. The macular organs, utricle and saccule, respond to translational head motion and convey the gravitational force vector^14^. Anatomically, the utricle and the saccule together form the vestibule. The cochlea is a separate inner ear organ for hearing. It consists of the scala vestibuli, the scala tympani and the cochlear duct (with hair cells). These structures spiral 2.57 ± 0.28 times from the base to the cochlear cupula, or apex of the cochlea^13^.

Segmenting these sub-structures in the inner ear and quantifying their anatomic dimensions of is a crucial step for morphology-based studies in numerous domains. In neurology and clinical neuroscience, physiological and morphological alterations of the cochlea can result in hearing deficits or loss, whereas affections of the semicircular canals or otoliths can lead to vertigo and balance disorders^15,16^. In anthropology, the labyrinthine morphology of the inner ear carries functional and phylogenetic parameters that can be tracked over the course of mammalian evolution^17^, e.g. by comparing ancient Egyptian mummies to contemporary anatomy^18^. In developmental neurobiology, models of the inner ear labyrinth^19^ and the cochlea^11,17,20,21^ lie at the basis of comparative neuroanatomy and cross-species studies. In neurosurgery and otorhinolaryngology, an accurate and patient-specific shape model of the cochlea is crucial for neuronavigation of cochlear electrode implantation and simulation^22–24^, or for post-operative assessment of surgical trauma^13^. One of the most accurate, versatile and robust way to anatomically delineate and demarcate individual brain structures is to segment these structures based on a reference template and corresponding atlas^25^. These standard, or reference template images represent a shape-based average from a normative cohort, with highly resolved anatomical structures in their mean location. Image templates and structural atlases are also fundamental tools for statistical shape analysis^8,26^ and group-based image analysis^27^.

Numerous imaging studies have accurately quantified the inner ear morphology, but few have created the type of re-usable template and atlas necessary for automatic segmentation of new individual data. Even less are based on non-invasive MRI imaging. The majority of inner ear analyses currently available are based on ex-vivo micro-CT as the imaging modality^28–30^, a few also providing cyto-architecture scans based on synchrotron radiation^31^, ex-vivo ultra-high-field MRI^32^, or light- and electron microscopy^33^. These images provide highly precise measures of inner ear dimensions given their high image resolutions, some on the order of histological images^31^. Several other studies have reported detailed measurements of the inner ear’s membraneous structures^34^, including the semicircular canals, based on ex-vivo micro-CT^20, 30, 35^ and multi-detector CT^36^. Most of these studies do not provide the template and atlas material for application to new individuals or tissue samples. The only study we are aware of that does, requires ex-vivo preparation of inner ear sections and micro-CT images to apply the atlas^11^. However, due in part to the rarity of such imaging devices, it is not straightforward to reproduce these measures in more conventional in-vivo imaging studies.

Recently, MRI-based visualization of the inner ear, with^37, 38^ or without Gadolinium-based contrast agents^39–43^ has become a more popular method for imaging the inner ear structures in-vivo. It been used for the detection of endolymphatic hydrops^41^ and for diagnosis of sensorineural hearing loss associated with inner ear lesions^44^. In-vivo MRI has also been shown to be accurate enough to determine cochlear duct length in patients undergoing cochlear implantation^45^. Furthermore, advances in MR sequences have improved the image quality of the inner ear structures. Sequences, such as *Constructive Interference in Steady State* (CISS) imaging, with low susceptibility and a high contrast-to-noise ratio, have been used to image the inner ear for the detection of leakage of labyrinthine and cerebrospinal fluids^46^.

In this work, we combine recent developments in MRI in-vivo imaging of the inner ear, modern neuroimaging methodology, and highly precise micro-CT based measurements to create IE-Map, a novel template and atlas of the human inner ear and its substructures. IE-Map is based on non-invasive in-vivo MRI sequences acquired from 63 subjects (126 inner ears). We leveraged the signal and contrast-to-noise advantages of multiple MRI sequences (T2-, CISS-, and T2-weighted sequences), all without invasive contrast agents or radiation, to create a an atlas that can be used to segment new MRI images of patients and subjects with MRI images in these contrasts. The template morphology was validated in several ways. First, we computed a novel inner ear template from 21 publicly available inner CT and micro-CT volumes, and measured the overlap and distances of inner ear surfaces between micro-CT and our T2 and CISS templates. Second, we validated our template and atlas annotations by measuring various inner ear dimensions and comparing them against precise measurements in micro-CT literature. We annotated our template with a set of 94 landmarks of the bony labyrinth commonly found in literature and with surface meshes of 29 anatomical sub-regions of the inner ear. Quantitative size and volume information of the inner ear substructures can then be extracted for each individual and side. To demonstrate this application of IE-Map we posed three questions about the population statistics of the inner ear. 1. Does head size, as measured by intracranial volume, influence labyrinth measurements? 2. Does gender have an effect in inner ear structure size? 3. Does ear laterality play a role in inner ear dimensions? The proposed template and atlas material in IE-Map is comprehensive, applicable, accessible, and generalizable. It thus has the potential to form the basis for future non-invasive neuroimaging studies of the inner ear under the umbrella term computational neuro-otology.

## Results

### Inner ear template and atlas

We applied a state-of-the-art two-step template building approach, first of a full-brain template, and second of a localized high-resolution inner ear template (cf. Fig. 4). Both templates were successfully reconstructed using algorithms provided by the ANTs toolkit^27^. The full-brain template (T2- and T1-weighted MRI) allowed for a fully automatic localization of both inner ears and a re-orientation of the axial slice to Reid’s standard plane for all subjects and inner ears. Consequently, the high-resolution (0.2 mm isotropic) templates in IE-Map (T2-, CISS-, and T1-weighted MRI) are aligned to Reid’s standard plane, roughly with the horizontal semicircular canal (SCC) in the axial plane.

Volume renderings of MRI templates can be seen in Fig. 1a (T2-weighted), Fig. 1b (CISS-weighted) and Fig. 1c (T1-weighted). The rendering transfer function was set as a linear ramp between the minimum and maximum voxel intensity for each template, no manual adjustments other than a shift of the centre intensity (T1: 1.2; T2: 1.5; CISS: 2.3) was necessary to achieve the visibility of inner ear structures as displayed. As noticeable from the rendering, the inner structures of semicircular canals, vestibule and cochlea are clearly visible and appear morphologically correct. Fine anatomical details and inter-template differences become apparent as well, e.g. a clear separation of scala tympani and vestibuli in the cochlea between the T2-weighted template (Fig. 1d) and the CISS-weighted template (Fig. 1e). The volume rendering does not suffer from noise artefacts observed in single-subject volumes, which we attribute to the fact that the multivariate templates on average have a 1.7 times higher signal-to-noise ratio (SNR) than the mean-SNR in single-subject volumes (see Table 1). SNR is approximated as the mean voxel intensity divided by the standard deviation of voxel intensities inside the inner ear ROI^47^.

**Figure 1.**
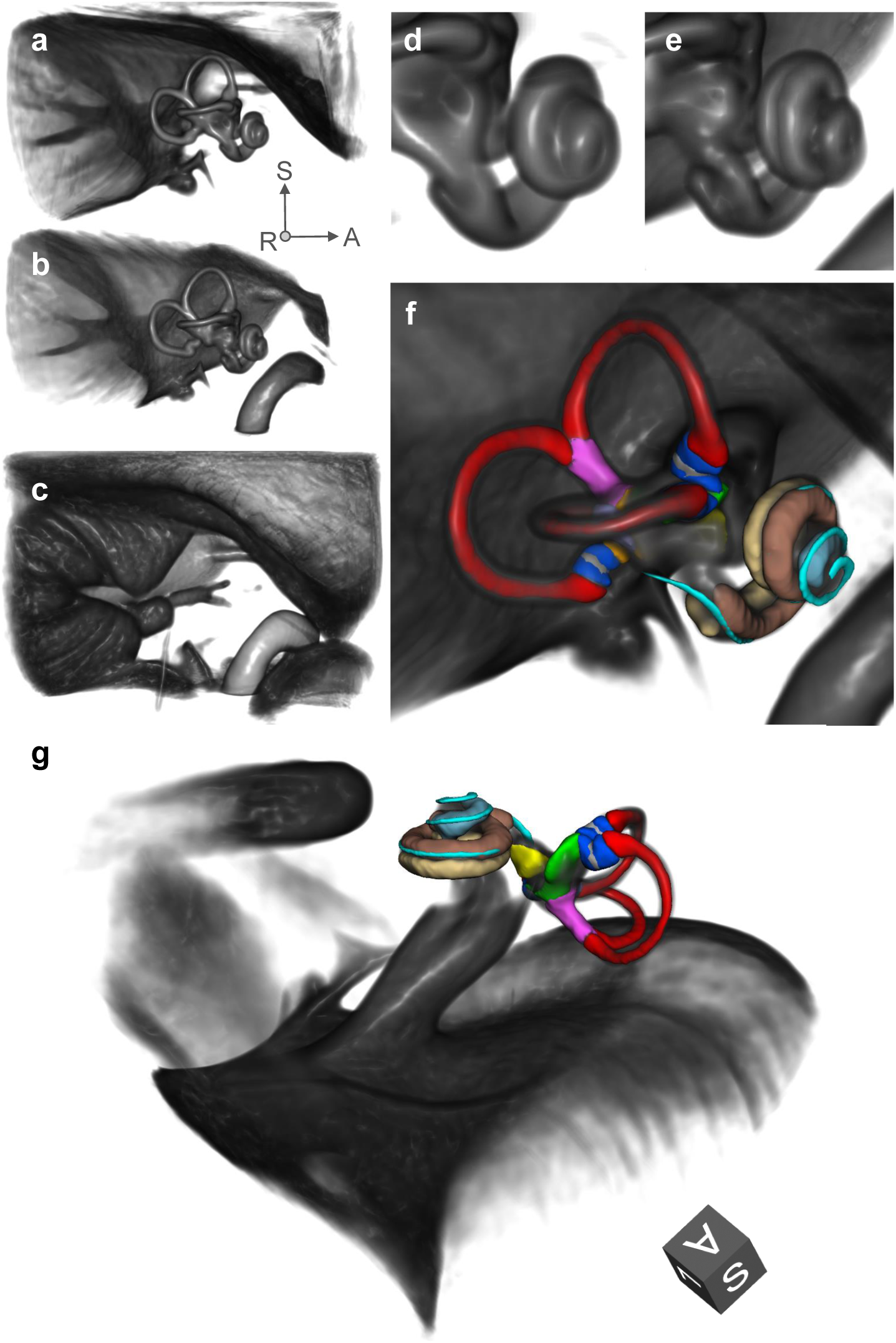
Volume renderings of inner ear templates and atlas. ROI size: 4×4×3 cm; resolution: 0.2 mm isotropic; viewpoint: right-lateral view onto right inner ear. Panels: a. T2 template; b. CISS template; c. T1 template; d. Closeup of cochlea in T2 template; e. Closeup of cochlea in CISS template, with a clear separation of scala tympani and vestibuli; f. Atlas surface meshes on CISS template volume rendering. g. Same as f, with a different perspective focussing on the vestibule. Colours and regions: red: semicircular canals; blue: ampullae; pink: common crus; grey: cupula walls; green: utricle; yellow: saccule; brown: scala vestibuli; beige: scala tympani; light blue: cochlear cupula; cyan: cochlear duct. The coordinate system arrows (panels a-f) and cube (panel g) indicate the anterior (A), superior (S) and left/right (L/R) directions.

**Table 1.**
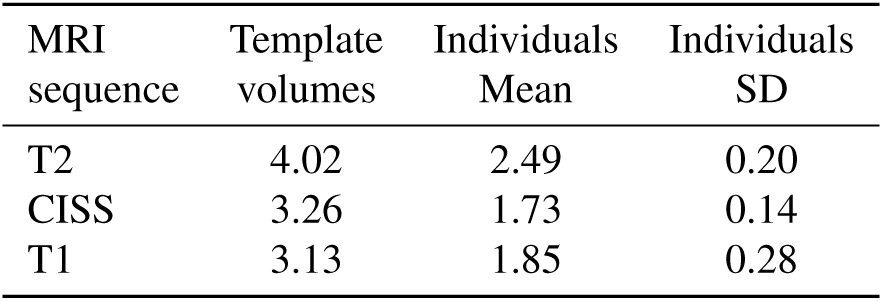
Signal-to-noise ratios in multivariate MRI templates are on average 1.7 times higher than in individuals’ volumes, resulting in improved visibility of structural details. Rendering can be performed nearly free of noise (cf. Figure 1), and fine-grained structures like the separation of cochlear scalae become visible in templates that are sometimes not even clearly discernible in individual volumes (cf. Figure 1, panel e).

| MRI sequence | Template volumes | Individuals Mean | Individuals SD |
| --- | --- | --- | --- |
| T2 | 4.02 | 2.49 | 0.20 |
| CISS | 3.26 | 1.73 | 0.14 |
| T1 | 3.13 | 1.85 | 0.28 |

### Cohort measurements and atlas validation

We first validated the inner ear morphology in our IE-Map templates by comparing it against CT and micro-CT imaging. Since our template represents an average morphology from 126 inner ears, we required a CT/micro-CT counterpart template which also represented an average morphology from a large cohort. To the best of our knowledge, such a template has not been computed yet or publicly shared. However, a recently published open-domain dataset offered co-registered, high-resolution clinical CT (0.15 mm isotropic) and micro-CT (0.06 mm isotropic) image volumes of 23 post-mortem preparations of the inner ear and bony canals^48^. We therefore utilized 21 of these volumes to create a novel morphological average template (0.06 mm isotropic) of the inner ear with multivariate CT/micro-CT, analogous to the ANTs based template building of IE-Map^27^. The CT/micro-CT template was only rigidly registered to IE-Map to account for global positioning and rotation, without any scaling or deformation of the CT morphology. An overlap of both templates can be seen in Fig. 2. Both templates have a remarkably high overlap, both in terms of Dice overlap coefficient^49^ (Dice overlap CT/CISS: 0.830, Dice overlap CT/T2: 0.861), as well as in terms of low surface average distances on the order of 0.1-0.2 mm (CT/CISS: min −1.36 mm, max 0.34 mm, mean −0.13 mm, 5th percentile −0.52 mm, 95th percentile 0.12 mm; CT/T2: min −0.94 mm, max 0.72 mm, mean 0.11 mm, 5th percentile −0.43 mm, 95th percentile 0.45 mm).

**Figure 2.**
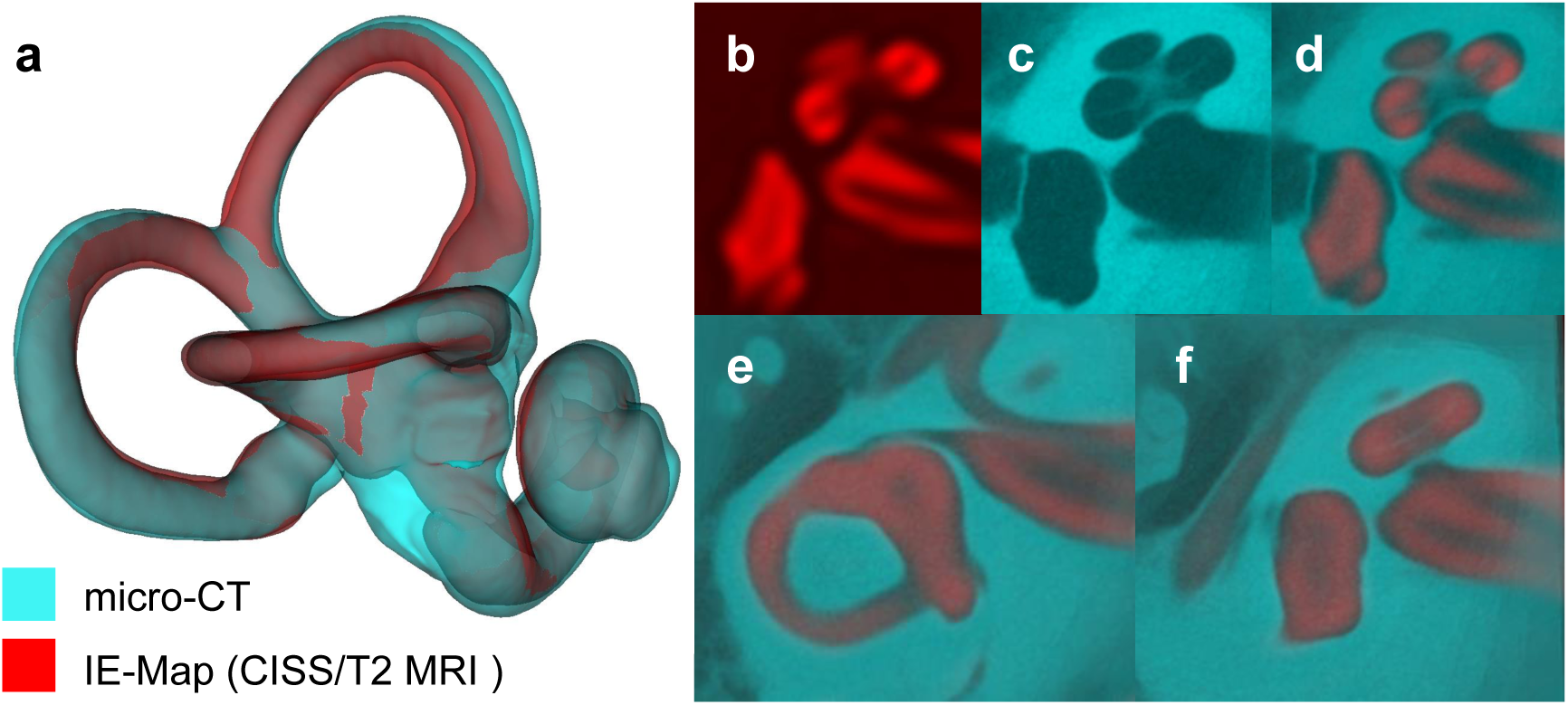
Validation of IE-Map morphology (red colormap) against an inner ear template built from 21 post-mortem CT and micro-CT scans (cyan colormap). Panels: a. 3D surface models of the inner ear in CISS MRI and micro-CT, showing a high overlap of cochlear and vestibular dimensions and morphology. Note that both templates represent the average morphologies from two independent cohorts, and that templates were only rigidly registered to account for global position and rotation of overlap. b.-d. Axial slice through the cochlea in CISS, micro-CT and a semi-transparent overlay. Note that the gap between scala tympani and vestibuli in the CISS volume coincides with the spiral lamina visible in the micro-CT template. e.-f. Morphological congruency of the lateral SCC (e.) and the vestibulum and basal cochlear turn (f.) in T2 MRI and micro-CT.

Surface models of the anatomic regions were further annotated in the template space of IE-Map and are visualized in Fig. 1 (panels f and g). Surface meshes of semi-circular canals, crus and ampullae were manually fine-tuned, after transfer from a micro-CT atlas^11^, cochlear scalae and cupula were segmented semi-manually based on contrasts in the CISS template, while utricle, saccule and cochlear duct were augmented from the micro-CT atlas through non-linear transformation.

Landmark annotations are visualized in Fig. 5 (panels a-d). The definition of landmark positions and distances between them followed conventions from micro-CT literature, details are given in Table 4. Based on landmark annotations, the characterization of the three semicircular canals (see Table 2) was compared to previously reported results using micro-CT imaging of ex-vivo specimens^35^. In general, our atlas produced slightly larger values than reported in literature, however, the error margin never exceeded the original image resolution of T1- and T2-weighted MRI (0.75 mm). Overall, the sub-voxel accurate distance measures demonstrate that the computed template has morphometric properties that are in line with previous ex-vivo micro-CT studies. The same applies to landmarks of the common crus and cochlea (see Table 3), which all corresponded to values in literature within a margin of less than 0.5 mm.

**Table 2.** Dimensions of the anterior (Ant), posterior (Post) and lateral (Lat) semicircular canals (SCC). SD indicates standard deviation. Unit: [mm]. * indicates non-normal distribution (Kolmogorov-Smirnov test with Lilliefors correction, *p <* 0.05).

| SCC | Dimension | Mean | SD | Mean<br>35 | SD<br>35 | ΔMean | p-val<br>L vs. R |
| --- | --- | --- | --- | --- | --- | --- | --- |
| Ant | Height | 6.4 | 0.5 | 6.5 | 0.5 | 0.1 | 0.074 |
|  | Width | 8.2 | 0.5 | 8.0 | 0.5 | 0.2 | 0.943 |
|  | Radius | 3.7 | 0.2 | 3.6 | 0.2 | 0.1 | 0.088 |
|  | Internal width 1 | 1.8 | 0.1 | 1.2 | 0.2 | 0.6 | 0.995 |
|  | Internal width 2 | 1.4 | 0.2 | 0.9 | 0.2 | 0.5 | 0.779 |
|  | Internal width 3 | 1.6 | 0.2 | 1.2 | 0.1 | 0.4 | *0.995 |
|  | Internal height 1 | 1.5 | 0.2 | 1.0 | 0.1 | 0.5 | *0.571 |
|  | Internal height 2 | 1.3 | 0.2 | 0.7 | 0.1 | 0.6 | 0.943 |
|  | Internal height 3 | 1.6 | 0.2 | 0.9 | 0.1 | 0.7 | 0.327 |
| Post | Height | 7.0 | 0.6 | 6.7 | 0.6 | 0.3 | 0.921 |
|  | Width | 7.7 | 0.6 | 7.7 | 0.5 | 0.0 | 0.695 |
|  | Radius | 3.7 | 0.3 | 3.6 | 0.2 | 0.1 | 0.623 |
|  | Internal width 1 | 1.8 | 0.1 | 1.4 | 0.2 | 0.4 | 0.695 |
|  | Internal width 2 | 1.7 | 0.2 | 1.5 | 0.2 | 0.2 | 0.995 |
|  | Internal width 3 | 1.6 | 0.2 | 1.4 | 0.2 | 0.2 | *0.995 |
|  | Internal height 1 | 1.4 | 0.1 | 1.0 | 0.1 | 0.4 | 0.779 |
|  | Internal height 2 | 1.3 | 0.2 | 1.0 | 0.1 | 0.3 | *0.995 |
|  | Internal height 3 | 1.4 | 0.2 | 0.9 | 0.1 | 0.5 | *0.995 |
| Lat | Height | 5.0 | 0.4 | 4.9 | 0.6 | 0.1 | 0.088 |
|  | Width | 6.6 | 0.4 | 6.5 | 0.7 | 0.1 | 0.571 |
|  | Radius | 2.9 | 0.2 | 2.9 | 0.3 | 0.0 | 0.088 |
|  | Internal width 1 | 1.8 | 0.2 | 2.1 | 0.3 | 0.3 | *0.716 |
|  | Internal width 2 | 1.7 | 0.2 | 1.7 | 0.3 | 0.0 | *0.995 |
|  | Internal width 3 | 2.2 | 0.1 | 1.7 | 0.2 | 0.5 | 0.164 |
|  | Internal height 1 | 1.8 | 0.2 | 1.2 | 0.2 | 0.6 | 0.995 |
|  | Internal height 2 | 1.4 | 0.1 | 0.8 | 0.1 | 0.6 | *0.954 |
|  | Internal height 3 | 1.7 | 0.1 | 1.2 | 0.3 | 0.5 | *0.530 |

Inner ear volume measurements were also largely in line with related literature (see Table 3), with a notable difference of almost +50 mm^3^ in total inner ear volume. Deviations from mean volumes reported in^50^ can be attributed to simplified geometric approximations made in that study. For example, authors in^50^ modelled the ampullae as spheres with 2 mm diameter (i.e. 4.2 mm^3^; ours: 3.54/2.27/3.59 mm^3^ for ant/post/lat ampulla), saccule as a hemisphere of 2 mm diameter (i.e. 2.1 mm^3^; ours: 2.6 mm^3^), utricle as an irregular oval averaging 1.35 mm in diameter and 5.5 mm in length (i.e. 7.9 mm^3^; ours: 8.8 mm^3^), and the cochlea as a cone with 8 mm diameter and 5 mm height (i.e. 83.8 mm^3^; ours: 94.4 mm^3^). The larger volume measurements of the cochlea and the total inner ear region indicate an increased estimate of the inner ear region by our T2 iso-surface, by 11% and 12%, respectively.

**Table 3.** Inner ear distances (in mm) and volumes (in mm^3^) in IE-Map (“our”), compared to micro-CT based measurements in literature (“lit.”). (n.r. = not reported).

| Distance / Volume | Mean (our) | SD (our) | Mean (lit.) | SD (lit.) | ΔMean | Reference |
| --- | --- | --- | --- | --- | --- | --- |
| Common crus length | 2.3 | 0.2 | 2.0 | 0.5 | 0.3 | 35 |
| Common crus width 1 | 2.2 | 0.2 | 1.6 | 0.3 | 0.6 | 35 |
| Common crus width 2 | 1.9 | 0.2 | 1.5 | 0.3 | 0.4 | 35 |
| Common crus width 3 | 2.3 | 0.2 | 1.9 | 0.4 | 0.4 | 35 |
| Cochlea height coronal | 5.58 | 0.26 | 5.31 | 0.52 | 0.27 | 51 |
|  | 5.58 | 0.26 | 5.11 | 0.30 | 0.47 | 52 |
| Cochlea height oblique | 3.56 | 0.22 | 3.59 | 0.12 | 0.03 | 53 |
| Cochlea length | 8.92 | 0.34 | 9.32 | 0.53 | 0.40 | 54 |
|  | 8.92 | 0.34 | 8.84 | 0.29 | 0.08 | 53 |
| Cochlea width | 6.78 | 0.31 | 6.30 | 0.38 | 0.48 | 53 |
| Vestibule length | 5.39 | 0.26 | 5.45 | 0.54 | 0.06 | 55 |
| Vestibule width | 3.08 | 0.17 | 3.20 | 0.39 | 0.12 | 55 |
| Inner ear length total | 17.59 | 0.76 | 17.13 | 0.64 | 0.45 | 56 |
| Vol. ampulla (ant) | 3.54 | 0.43 | 4.20 | n.r. | 0.66 | 50 |
| Vol. ampulla (post) | 2.27 | 0.27 | 4.20 | n.r. | 1.93 | 50 |
| Vol. ampulla (lat) | 3.49 | 0.45 | 4.20 | n.r. | 0.71 | 50 |
| Vol. saccule | 2.58 | 0.41 | 2.10 / 2.10 | n.r. | 0.48 | 50, 57 |
| Vol. utricle | 8.78 | 1.12 | 7.90 / 8.20 | n.r. | 0.88 / 0.58 | 50, 57 |
| Vol. cochlea | 94.42 | 9.41 | 83.80 | n.r. | 10.62 | 50 |
| Vol. inner ear total | 269.34 | 24.67 | 221.47 | 24.29 | 47.87 | 56 |

**Table 4.** Definitions of inner ear measures, adopted from indicated source references into IE-map (where applicable; MPR = Multi-planar reconstruction, i.e. an arbitrary slice angulation through the voxel volume.)

| Measure | Definition | Source |
| --- | --- | --- |
| SCC width | [Visual guideline without explicit instructions] | 35 |
| SCC height | [Visual guideline without explicit instructions] | 35 |
| SCC radius | $SCC\ radius = 0.5 (SCC\ height + SCC\ width)/2$ | 35 |
| SCC inner width and height | Internal widths and heights of the three SCCs were measured perpendicularly, at three points from the distal common crus to the proximal common crus. | 35 |
| Common crus length | [Visual guideline without explicit instructions] | 35 |
| Common crus width | Common crus width was measured at three points: superior, middle, and inferior. | 35 |
| Cochlea height coronal | Maximum height that included the basal and upper turn, measured on a coronal cut. | 51 |
| Cochlea height coronal | Maximum height measured perpendicularly to the oval window. | 52 |
| Cochlea height oblique | Slice view: MPR slice aligned to an oblique coronal view perpendicular to the cochlear basal turn; Landmarks: cochlea height is defined as the length of the line from the cochlear nerve foramen to the cochlear apex. | 53 |
| Cochlea length | Slice view defined in<sup>53,54</sup>: basal turn of the cochlea viewed by minimum intensity projection using a 1.0-mm layer; Slice view defined here: least-squares plane fit through seven equidistant points distributed across the center spiral through the basal turn (fitting landmarks shipped with open-source material); Landmarks: largest distance from the round window to the lateral wall through the modiolar axis. | 53,54 |
| Cochlea width | Length of the line passing through the modiolar axis, perpendicular to the cochlea length line, and intersecting with the upper and lower edge of cochlea. | 53,54 |
| Vestibule length | Maximal longitudinal diameter | 55 |
| Vestibule width | Maximal transverse diameter | 55 |
| Inner ear length total | Line distance from the most posterior point of the posterior SSC to the most anterior point of the cochlea. | 56 |
| Vol. ampulla (ant) | Volume of registered surface mesh from a reference micro-CT atlas. | 11 |
| Vol. ampulla (post) | Volume of registered surface mesh from a reference micro-CT atlas. | 11 |
| Vol. ampulla (lat) | Volume of registered surface mesh from a reference micro-CT atlas. | 11 |
| Vol. saccule | Volume of registered surface mesh from a reference micro-CT atlas. | 11 |
| Vol. utricle | Volume of registered surface mesh from a reference micro-CT atlas. | 11 |
| Vol. inner ear total | Total volume of inner ear structures in the T2-weighted template (Otsu-threshold iso-surface) | n.a. |
| Vol. cochlea | Cochlea volume int T2-weighted template (Otsu-threshold iso-surface) | n.a. |

### Cohort analysis and statistics

As detailed in Table 2, paired univariate testing with multiple testing correction yielded no significant differences regarding sidedness in any of the semicircular canals measures (height, width, radius, as well as internal height and width measured at three distinct points from the distal to the common crus). We further investigated the major inner ear dimensions, i.e. total inner ear length, total inner ear volume, oblique cochlea height, and radii of the three semicircular canals. First, we found all these values to be normally distributed (KS-test with Lilliefors correction, *p* > 0.05). Second, we investigated a possible correlation of selected inner ear measures with brain size measured by total intracranial volume (TIV). We found the total inner ear volume and length, as well as the radii of the anterior and posterior semicircular canals (SCC) to be moderately and significantly correlated with brain size, while the oblique cochlea height and the radius of the lateral SCC were not significantly correlated (cf. Fig. 3). Third, we investigated whether there is a significant difference between male and female subjects in our cohort, on the main and most robust inner ear measures, i.e. the total inner ear length and total inner ear volume. Since TIV was found to be a confounding variable for these measures, we performed a One-way ANCOVA, controlling for TIV as a covariate. Our ANCOVA tests yielded no significant differences for the investigated inner ear measures (total inner ear length: *F*_1_,_62_ = 1.82, *p* > 0.05; total inner ear volume: *F*_162_ = 3.02, p > 0.05). In other words, after accounting for overall brain size, gender difference cannot explain residual differences in measured inner ear characteristics.

**Figure 3.**
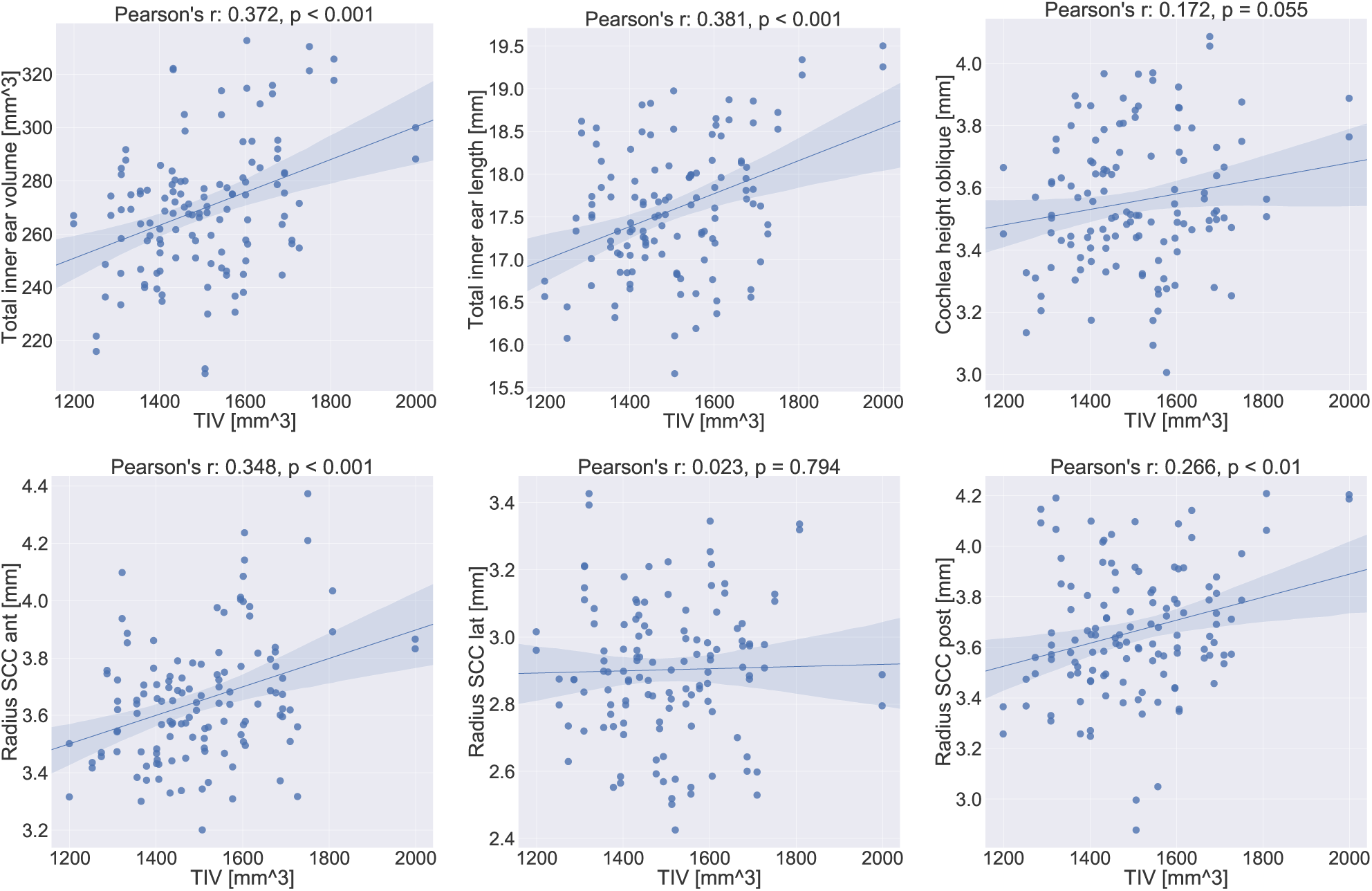
Correlation plots for inner ear measurements with total intracranial volume (TIV). Linear correlation values (Pearson’s r) and corresponding p-values are indicated in the subplot titles. The 95% confidence interval for the regression estimate is drawn as a translucent band around the regression line. Total inner ear volume and length, as well as the radii of the anterior and posterior semicircular canals (SCC) are linearly correlated with head size. The oblique cochlea height might well be while the radius of the lateral SCC appears to not be correlated at all with TIV. The results indicate the importance of TIV for future quantitative assessments between cohorts.

## Discussion

In this work, we presented a human inner ear atlas derived from multi-sequence magnetic resonance imaging in-vivo and non-invasively. To the best of our knowledge, this is the first multi-sequence MRI template and atlas combination of the human labyrinth, covering vestibular and auditory structures, and the first time it has been constructed from a sizable cohort (>50 subjects). The segmentation and annotation of templates in our work benefited predominantly from the SNR of the source whole-head T2 images in combination with the high spatial resolution of the CISS sequence. The data analyses resulted in an inner ear atlas uniquely depicting the vestibule, the three semicircular canals and their ampullae, and the cochlea with its two scalae (scala vestibuli, scala tympani) and the cochlear cupula. Using registration-based augmentation of further structures from the micro-CT atlas from David et al.^11^, we can further visualize the position and shape of the cochlear duct, utricle and saccule. The TIV was found to be a relevant covariate for measurements of the labyrinth and its substructures. After controlling for TIV as a covariate, no gender or laterality effects were found.

Our reported region volumes and landmark distances were in line with the published literature. We were able to reproduce landmark locations and distances with the help of detailed 3D visualizations of landmark locations in several works^35,53^. We followed these closely in our work, and visualized them for reference in a similar fashion as in^35^ (cf. Fig. 5). Earlier descriptions of landmark placement were often based on 2D volume planes, which can be more difficult to understand and reproduce. For example, the cochlear height was previously defined either in the coronal image plane^51,52^, or using an oblique volume cut through the centre of the cochlear basal turn and the cochlear apex^53^. While the former depends on the orientation of the inner ear anatomy with respect to the coronal plane, the latter is uniquely defined in 3D space. Consequently, our measurements of the cochlear height deviate more from the measurements reported in either^52^ or^51^, but is almost identical to the ones in^53^. Further improvement is difficult to achieve without a significantly higher voxel resolution in the source material, which would imply a higher MRI field strength. In any case, an exact replication of the values reported in the literature is not useful at this point because i) we relied on a young healthy cohort, while many of the literature we compared our landmarks to included pathologies^51,52,58^ ii) we relied on a single-rater landmark placement, and the related works did not consider intra- or inter-rater variability in landmark localization, and iii) it is not yet clear how comparable distance measures from MRI are to those from micro-CT.

Overall, no other publicly available template or atlas of the inner ear has assembled and published a comprehensive list of inner ear landmarks for distance measurements. It should be noted that the literature-oriented landmark annotation in our work was time-consuming and non-trivial. By publicly providing these landmarks along with our templates and segmentations, we hope to provide a reproducible and consistent basis for such measurements in future studies as well as to pave the road for population-based standard values. We recently presented another inner ear MRI template^59^, which is complementary to this work, with two major differences. That work uses intravenous Gadolinium contrast enhanced MRI, and the reconstruction of two separate templates for the left and right side. In this work, we present a fully non-invasive counterpart which further covers the use-case of inner ear imaging without contrast agent. We also extended our methodology to reconstruct a single template which is unbiased to the side, which allowed us to analyze potential laterality confounds for the first time. Our novel atlas now contains detailed labels for sub-structures of the semicircular duct system, vestibule and cochlea. These were obtained through manual labelling and registration of an atlas from micro-CT^11^. We validated the template and atlas by performing a morphometric analysis of the semicircular canals, vestibule and cochlea, and by comparing anatomical measurements to established measures from micro-CT literature. We further validated the template by reconstructing a novel CT/micro-CT template and ensuring morphological similarity through overlap and surface distance measures.

It is important to note that because of our inner ear template comes from, and exists in multiple MR sequence intensities, it provide a high level of flexibility for using this atlas. Future studies based on T1-, T2-weighted data and CISS-sequences can directly utilize our templates through intra-modal atlas registration (in a single- or multivariate approach). Other structural MRI sequences (with or without contrast), MRI paradigms (e.g. diffusion-weighted MRI) or even other modalities (e.g. CT, micro-CT) can be registered and utilized with our atlas, either through multi-modal image similarity functions (e.g. normalized mutual information), or directly through the CT/micro-CT templates we created during our validation step and which are included in IE-Map. Together, the T1/T2/CISS/CT/micro-CT templates cover a wide range of complementary image appearances for inner ear tissues.

Our atlas deviates slightly from the existing literature, in particular regarding volumetric measures. For example, the overall inner ear volume appears to be marginally over-estimated in our template. This may be due to partial volume effects and signal blurring after template building. It is likely in part due to a slightly different definition of the inner ear surface contour in IE-Map compared to literature. This is supported by the fact we built the MRI and CT/micro-CT templates in a similar fashion; surfaces in MRI and CT were extracted based purely on image-based information (Otsu thresholding) with minimal manual post-processing. The surfaces in both modalities are morphologically very similar, with high overlap (Dice coefficient > 0.85 for T2) and low surface distances (0.1-0.2 mm on average). Other volumetric measures like the ampullae appear to be slightly under-estimated compared to the literature. As mentioned above, this is likely due to simplified geometric assumptions for anatomic shapes made by previous authors^50^. These simplifications were probably helpful in this pioneering study, which approximated 3D volumes through 2D cross-cuts. Materials like our 3D templates and surface meshes should allow for more accurate volumetric and thus more realistic measurements. Overall, we find it remarkable that the outer surface of the inner ear in the T2-weighted template (i.e. the Otsu-threshold iso-surface) seems to accurately represent the outer surface of bonystructures as measured by micro-CT, a result that was not expected prior to conducting the study.

Our atlas can be applied for localization, segmentation and sub-parcellations of the entire human inner ear on new MRI or micro-CT/CT images from participants that were not used in this study. By co-registering this data to the template, individual differences can be detected (e.g. of the anatomical structures itself or with respect to deviations from Reid’s plane). We believe that the atlas will also be helpful for surgical planning for the implantation of cochlear or vestibular electrodes^45^. Even though malformations were not used to build our atlas, image-based registration is often robust enough to compensate for a wide range of deformations, such that we expect the material to be useful for in-vivo detection and comparison of morphological abnormalities of the inner ear structures as well.

A limitation of our MRI templates in IE-Map is that our 63 subjects were only scanned with MRI, and not additionally with high-resolution micro-MRI and/or CT/micro-CT imaging. This would have allowed us to validate with a structural gold standard, and directly investigate MRI-specific phenomena such as distortions at air-tissue interfaces. However, our goal was to provide an in-vivo template of a normative control cohort with healthy young subjects. Micro-CT/-MRI both require ex-vivo preparation of post-mortem specimens, and high-resolution CT leads to high doses of radiation. Therefore, a direct validation of healthy control subjects in these imaging modalities is not possible from an ethical standpoint. Nonetheless, we validated our landmarks using micro-CT in two different ways, through direct comparison against landmark measurements in literature, and through the novel CT/micro-CT template we created. Our results indicate that our MRI templates in IE-Map reproduce the inner ear anatomy faithfully and that air-tissue interface artifacts do not play a significant role. A further limitation is that our results still do not resolve every fine detail of the inner ear, especially the auditory aspects, despite the high isotropic voxel resolution of 0.2 mm in the final MRI IE-Map templates. We were able to visualize the three major parts of the cochlea (the scala vestibuli, the scala tympani and the cochlear duct), but the results were still insufficient for e.g. a visualization of the organ of corti that lies inside the cochlear duct and houses the hair cells. The same holds true for the structure inside the ampullae: the cupulae. These structures are not ossified and it is far more complex to detect them with 3T MRI contrasts due to the tissue type, the achievable isotropic resolution, and mitigating head motion during data acquisition. Notably, the CT/micro-CT templates were also not able to clearly resolve these structures, neither in the source material, nor in the reconstructed template, possibly due to image noise and limited resolution.

There are natural limits to MRI resolution at 3T, and more fine-grained structures can only be resolved with ex-vivo microscopic imaging techniques. However, in this work we demonstrate the feasibility of a multimodal fusion of high-resolution templates, by providing the first multivariate MRI template set of the inner ear. It seems feasible and realistic to achieve further improvements to the resolution of a high-resolution T2-SPACE with coverage of the temporal bone alone and the CISS sequence in the target range of 0.4mm. A further limitation of the study is that, because the cohort’s mean age was 26 +/− 2.3 years, the template is only for individuals with a fully formed adult human skull. Changes due to age can therefore not be addressed with this template. However, we would not expect a change in bony structures in elderly patients with hearing or vestibular problems. Typically, a degeneration of soft tissue like hair cells in the organ of corti or the cochlear nerve lead to age related hearing loss. Further inner ear malformations reported previously^58^ are not represented by our atlas, since we only investigated MRI images of normal subjects. Nonetheless, as noted, malformations may well be detectable as significant structural deviations, through co-registration and overlay on our template on a subject-basis.

There are several ways in which the proposed inner ear atlas can be extended. First, the atlas can be augmented by additionally labelling nearby regions. The auditory and the two vestibular nerves appear as separable structures in the templates and are hence of particular interest, but were out of scope of this inaugural study. Once these nerves are included, the facial nerve may as well be segmented as it runs in close proximity of the vestibule. Similar to the inner ear micro-CT atlas^11^, other micro-CT atlases as in^29^ can help in identifying and accurately delineating nerves, vasculature and bones in our template space. For nerves, the T1-weighted template could become more relevant than in this study. In T1, nerve structures are not only visible (compared to the rest of the inner ear structures), but they have a complementary, hyper-intense appearance, while appearing hypo-intense in the T2 and CISS templates (see Figure 1, panel c). Adding the relative angles of the semicircular canals with respect to one another on a single-subject basis and analysing the coplanarity of functional pairs^60^, together with the orientation of the eye muscles can further increase the clinical relevance of our IE-Map atlas. Another next step is to develop an automatic segmentation method for prospective validation and application of the templates and atlas. In initial registration experiments, we have verified that segmentation of new subjects is possible using pair-wise single- and multivariate image registration methods, again provided by ANTs^27^. A fully automatic segmentation method would require the tool-chaining of left and right inner ear localization, template displacement, and local deformation to the inner ear structures of test subjects. The implementation and validation of such a toolkit is also left for future work. A further exciting avenue for improvement stems from the multi-modal fusion of our in-vivo template with micro-CT volumes from multiple subjects. In-vivo MRI may never resolve a similarly fine-grained structural information as micro-CT, but we have demonstrated the feasibility of geometric fusion through multivariate image registration and surface alignment. Eventually, this could allow for the construction of probabilistic label maps in micro-meter resolution, within the unbiased geometry of our template space.

In summary, it is our hope and aim that template and atlas can find application in numerous domains and disciplines studying neurootological health and disease. Potential applications include inner ear localization, segmentation and sub-parcellation, to quantitatively assess peripheral vestibular and auditory anatomy in group comparisons as well as over the course of longitudinal studies. By providing the templates, atlas segmentation, surface meshes and landmark annotations freely and publicly, we hope to provide clinicians and researchers in neurology, neuroscience and otorhinolaryngology with a new tool and robust basis as a starting point for computational neuro-otology, grounded in neuroimaging.

## Methods

### Subject cohort

The template was created from a cohort of 63 healthy, right-handed subjects (32 female, 31 male, age: 26 ± 2.3 years). To make the template and atlas representative and generally applicable to both left and right inner ears, both sides were treated independently, resulting in 126 inner ears in total that were used for template building. All subjects gave written informed consent regarding participation in this study. The local ethics committee of the Ludwig-Maximilians-Universität, Munich, approved the study in accordance with the latest revision of the Helsinki declaration from 2013.

### MR imaging and pre-processing

We acquired multi-modal, isotropic 3D MRI data (whole-head T1- and T2-weighted at 0.75 mm, CISS measurement of the labyrinth at 0.5 mm isotropic) in a clinical 3T scanner with a 64-channel head/neck coil (Siemens Skyra, Erlangen, Germany). The high-resolution T1-weighted MPRAGE sequence was collected with GRAPPA in an interleaved mode and an acceleration factor 2 (TR 2060ms, TI 1040 ms, TE 2.17 ms, flip angle 12 deg., FoV 240 mm, slice thickness 0.75 mm, 256 slices, A-P phase encoding, echo spacing 7.9 ms, bandwidth 230Hz/Px, prescan normalized). A T2-SPACE sequence with varying flip angles (TR 3200 ms, TE 560 ms, FoV 240 mm, slice thickness 0.75 mm, 256 slices, A-P phase encoding direction, GRAPPA acceleration factor 2, echo spacing 4.06 ms, bandwidth 625Hz/Px, prescan normalized) was collected for an optimized contrast between tissue types for the atlas generation, to detect brain pathologies and achieve whole-head coregistration for all data. In addition, we acquired an optimised CISS sequence for labyrinthography (TR = 8.56 ms, TE = 3.91 ms, flip angle = 50 deg., FoV = 150 mm, slice thickness= 0.50 mm, right-left phase encoding direction, sequential multi-slice mode, tune up shimming, activation of head coil elements 5-7 and the cranial neck coil elements only, bandwidth 460Hz/pixel, the average of two repetitions was built, total time of acquisition 13:40 minutes) for achieving the best possible contrast and resolution for inner ear structures.

Head motion was controlled by a dedicated head fixation device (®Pearltec Crania Adult, Schlieren, Switzerland). All MRI image data underwent stringent quality assessment before inclusion in the study, using a state-of-the-art MRI quality control toolkit (MRIQC)^61^. As pre-processing, we performed a correction for MRI field inhomogeneity using the N4 bias correction algorithm^62^. Further, we performed a rigid intra-subject registration of T1 and CISS volumes to the T2 volume, before proceeding to template building. Intra-subject registration was performed using Advanced Normalization Tools (ANTs), with normalized mutual information for multi-modal alignment^27^.

Total intracranial volume (TIV) as a potential covariate for head size was obtained via segmentation of the T1-weighted image with the CAT12 toolbox (version 12.6 release 1450, http://www.neuro.uni-jena.de/cat/) of the Structural Brain Mapping group (Christian Gaser, Jena University Hospital, Jena, Germany), a toolbox implemented in SPM12 (v7487, Statistical Parametric Mapping, Institute of Neurology, London, UK).

### Template building

Template building was performed in three steps: 1) full-brain template building and annotation, 2) axial re-orientation of all subjects to the Reid’s axial plane, and 3) final inner ear template building. The template building method utilized in steps 1 and 3 is a state-of-the-art algorithm provided by the ANTs toolkit^27^. It is designed for computation of a geodesic mean template that optimally represents the average anatomic shape from a set of input volumes. Critically, all ANTs components are capable of multivariate image registration, allowing us to fully leverage the multi-sequence appearance of the inner ear and its surrounding anatomy in T1, T2 and CISS MRI during template reconstruction. The high accuracy and robustness of ANTs registration and template normalization methods have been validated by demonstrating state-of-the-art performance in several internationally recognized medical image processing challenges^25, 63^.

In the first step, we computed a full-brain template for our cohort using T1- and T2-weighted MRI sequences. For the full-brain template, CISS volumes had to be omitted since they were acquired only within a narrow axial field-of-view (FOV) centred around the inner ear region. In the full-brain template, we manually annotated four landmarks for identification of the Reid’s axial plane, i.e. the locations of the infraorbital point and external auditory meatus in the left and right hemisphere^64^. We further placed two landmarks at the locations of the left and right inner ear, for a consistent localization of the inner ears across all subjects. This procedure is illustrated in Fig. 4.

**Figure 4.**
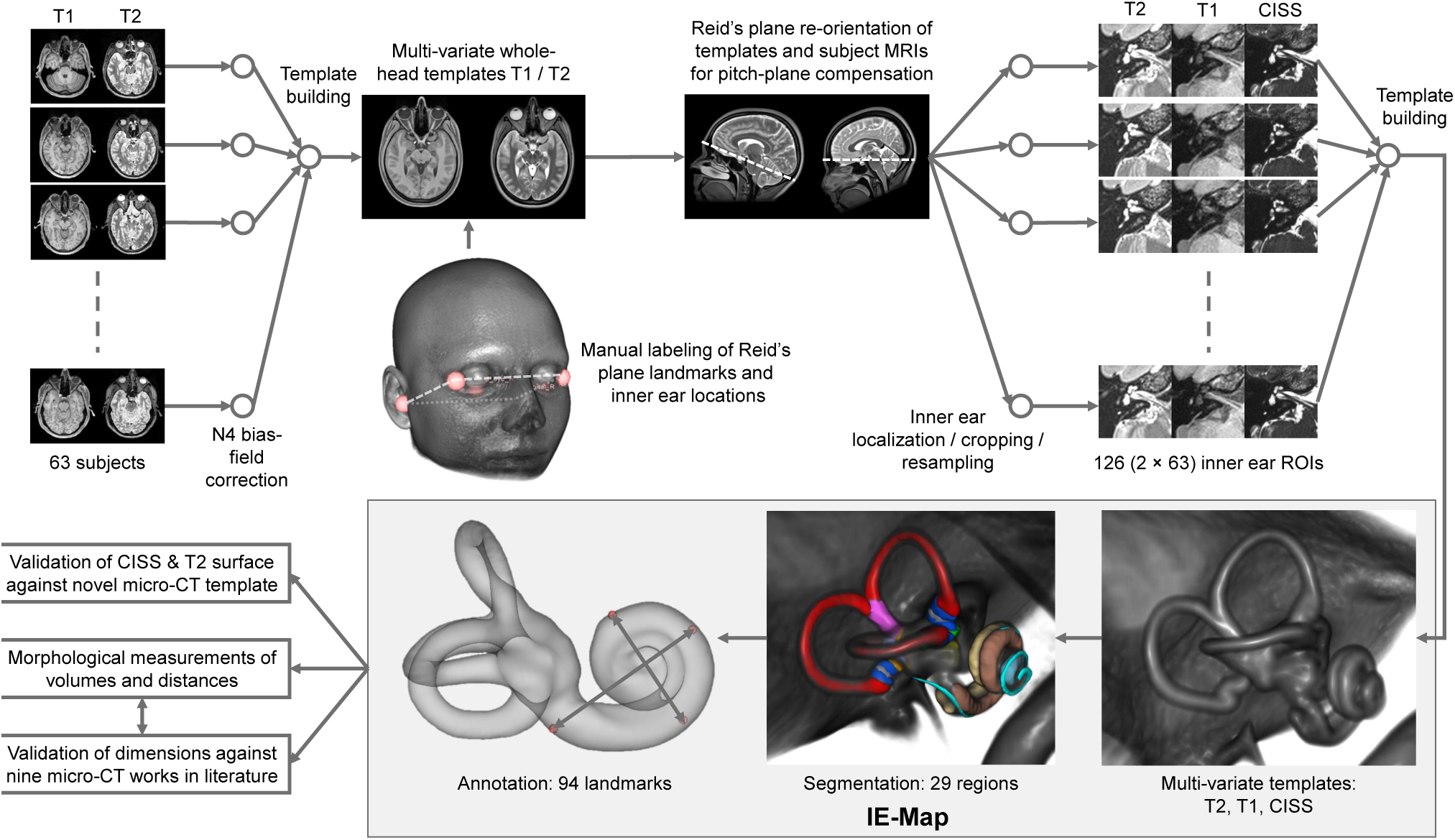
Workflow for reconstruction, annotation and validation of IE-Map. First, a full-brain template was reconstructed and annotated for re-orientation of the axial plane to Reid’s standard plane, and for localization of left/right inner ear region-of-interests (ROI). The rendered template head represents an average morphology of our cohort. Second, the inner ear template was reconstructed from 126 example multivariate inputs (T2, T1, CISS). The high-resolution (0.2 mm isotropic) template was annotated with landmarks, and segmented, partly through registration of a publicly available micro-CT atlas^11^. To validate the morphology, we compared inner ear surfaces from T2 and CISS templates to a novel micro-CT template, as well as landmark distances to nine micro-CT based works in literature. Templates, segmentation labelmaps, landmarks, and surface models together form IE-Map, which we publicly provide as open-access material.

In the second step, we re-oriented source volumes to Reid’s standard plane. This was a necessary pre-processing step for the inner ear template building (step 3), which took crops around the inner ear’s region-of-interest (ROI) as input. Pre-aligning these crops resulted in less out-of-border interpolation, i.e. less black margins and a better registration result at the ROI borders during template building. A side benefit of this step was that the lateral semicircular canal became approximately aligned with the axial plane, given the common assumption of coplanarity of the lateral semicircular canals with Reid’s standard plane^65–67^. The landmarks for Reid’s plane and inner ear FOV centres were transformed into the volumes of all input subjects beforehand, using a full-brain registration from template to individual spaces, followed by visual inspection for quality control. We then computed Reid’s plane parameters using least-squares fitting to the four landmarks, and re-oriented the axial plane in all volumes accordingly.

In the third step, we used the inner ear landmarks identified in step 2 to place region-of-interest (ROI) bounding boxes at the left and right inner ear. At these locations, the T2, CISS and T1 volumes were cropped (4×4×3 cm) and resampled (0.2 mm isotropic, bi-cubic B-spline interpolation) with a ROI bounding box that included all relevant inner ear structures as well as the vestibulocochlear nerve. As performed during pre-processing on the full brain volumes, we linearly registered the CISS and T1 crops to the T2 crop, to ensure a perfect alignment of fine structures of the inner ear across all MRI contrasts. Cropped volumes were centred and all left inner ears were mirrored with respect to the sagittal plane, to match the orientation of the right inner ear. This latter step ensured that the obtained template is unbiased with respect to afference laterality (sidedness) of the inner ear structures. The above steps resulted in 126 localized, multivariate MRI volumes, oriented to Reid’s plane and hence roughly oriented with the lateral semicircular canal in the axial plane. These volumes served as input to the multivariate template building algorithm, which computed three inner ear templates at an isotropic target resolution of 0.2 mm. The procedure and resulting template are illustrated in Fig. 4.

In a fourth step, we used publicly available CT and micro-CT volumes from 23 subjects^48^ to build a novel, multivariate CT/micro-CT template of the inner ear, which we used towards validating the morphology of our MRI templates. Since the source images were oriented arbitrarily in space, we first performed a rigid pre-alignment of CT/micro-CT volumes to IE-Map (i.e. to Reid’s plane orientation) through consecutive landmark registration and image-based refinement. Source images contained two artifacts which prohibited image-based registration, namely fixation pins, which were used to hold the post-mortem specimens in place inside of the CT/micro-CT scanners, as well as zero-voxel areas in micro-CT volumes due to co-registration and resampling into the CT reference frame. These areas were masked and filled with the average tissue intensity of the inner ear region, providing a tissue-neutral surrounding during registration. Subsequently, we utilized the same ANTs-based template building algorithm as for MRI templates in IE-Map, this time in a multivariate manner on paired CT/micro-CT volumes. We were able to include 21 out of 23 source images from^48^, since two volumes could not be pre-aligned with IE-Map in an image-based manner. The outer surface of the bony labyrinth was extracted from the resulting CT/micro-CT template via thresholding and manual post-processing in 3D Slicer^68^. The CT/micro-CT template was registered rigidly to IE-Map one last time to ensure structural alignment between both template spaces. It is important to note that both templates were built from independent cohorts and represent optimal structural average estimates of the inner ear anatomy in these two modalities. Both template geometries were registered only rigidly, i.e. without any linear or non-linear distortions that would affect the morphological measurements performed during validation.

### Atlas annotation

The multi-sequence template was segmented into anatomic regions in order to obtain an atlas of the inner ear. Segmentations were obtained in parts by transferring fine-grained surface meshes of the semicircular canals, vestibule and cochlea from *Ariadne*, a publicly available atlas and segmentation toolkit of the inner ear derived from ex-vivo micro-CT data^11^. From this toolbox, we leveraged micro-meter accurate surface meshes as delineations of inner ear structures, by registering them to our multivariate MRI template.

Due to the lack of an accompanying micro-CT intensity volume in the Ariadne toolkit, we performed a volumetric registration and geometric transfer through alignment of the entire inner ear’s outer surface. For our T2 template volume, we computed the outer iso-surface of the inner ear at an optimal intensity threshold obtained by Otsu’s method^69^, followed by manual removal of background structures that were obtained during thresholding. For the micro-CT data, we utilized the bony surface mesh provided in the *Ariadne* toolbox. Next, we voxelized both the T2 surface mesh and the micro-CT surface mesh of the inner ear at an isotropic resolution of 0.1 mm, and computed signed Euclidean distance transforms (SDT) for both meshes^70^. These SDT representations served as surrogate image volumes during deformable image registration, yielding a non-linear deformation field from Ariadne atlas space into our template space. All micro-CT surfaces including semicircular canals, vestibule, and cochlear ducts (i.e. scala vestibuli, scala tympani and cochlear duct) were then transferred onto our template volume and voxelized at the resolution of 0.2 mm. The resulting labelmap was manually post-processed, based on visual inspection, to ensure alignment of labels with the three intensity channels T2, T1 and CISS. Volumes of utricle, saccule and ampullae were estimated based on micro-CT meshes^11^ co-registered into our template.

The cochlea regions were pre-segmented by intensity-based thresholding of the CISS template. The threshold was set to obtain a clear separation of the scala tympani and vestibuli. Both regions were refined manually using basic image processing operations such as connected components, and surface smoothing. The cochlear duct is not directly visible in MRI templates, but was augmented in template space through non-linear transformation from the micro-CT atlas^11^ using the computed deformation fields.

The procedures described above were realized using various tools in the open-source software toolkits ANTs (deformable surface registration)^27^, 3D Slicer (segment editor effects: threshold, connected components, paint, scissors, margin)^68^ and SimpleITK (Euclidean SDT computation with SignedDanielssonDistanceMapImageFilter)^71^.

Using landmark annotation in 3D Slicer^68^, we further annotated multiple anatomic landmarks of the inner ear structures described in related literature. For the sake of reproducibility and annotation consistency, we placed all landmarks on the Otsu-threshold iso-surface of the T2 template volume. The landmark locations on the T2 iso-surface are visualized in detail in Fig. 5. Following guidelines and 3D visualizations in^35^, we annotated the three semicircular canals regarding width and height, as well as inner widths and heights measured at three positions (Figure 5, panels a and b). We also followed instructions in^35^ to compute the height and three width measures of the common crus. The height and width of the vestibule were described in^55^. The total inner ear length was described in^56^ (Figure 5, panel c). The placement of landmarks for measurement of cochlea width, length and height was described in^53^ (Figure 5, panel d). While cochlear height here was defined on an oblique 3D cut through the volume, earlier approaches defined cochlear height on the coronal plane^51,52^, which we also measured for comparison.

**Figure 5.**
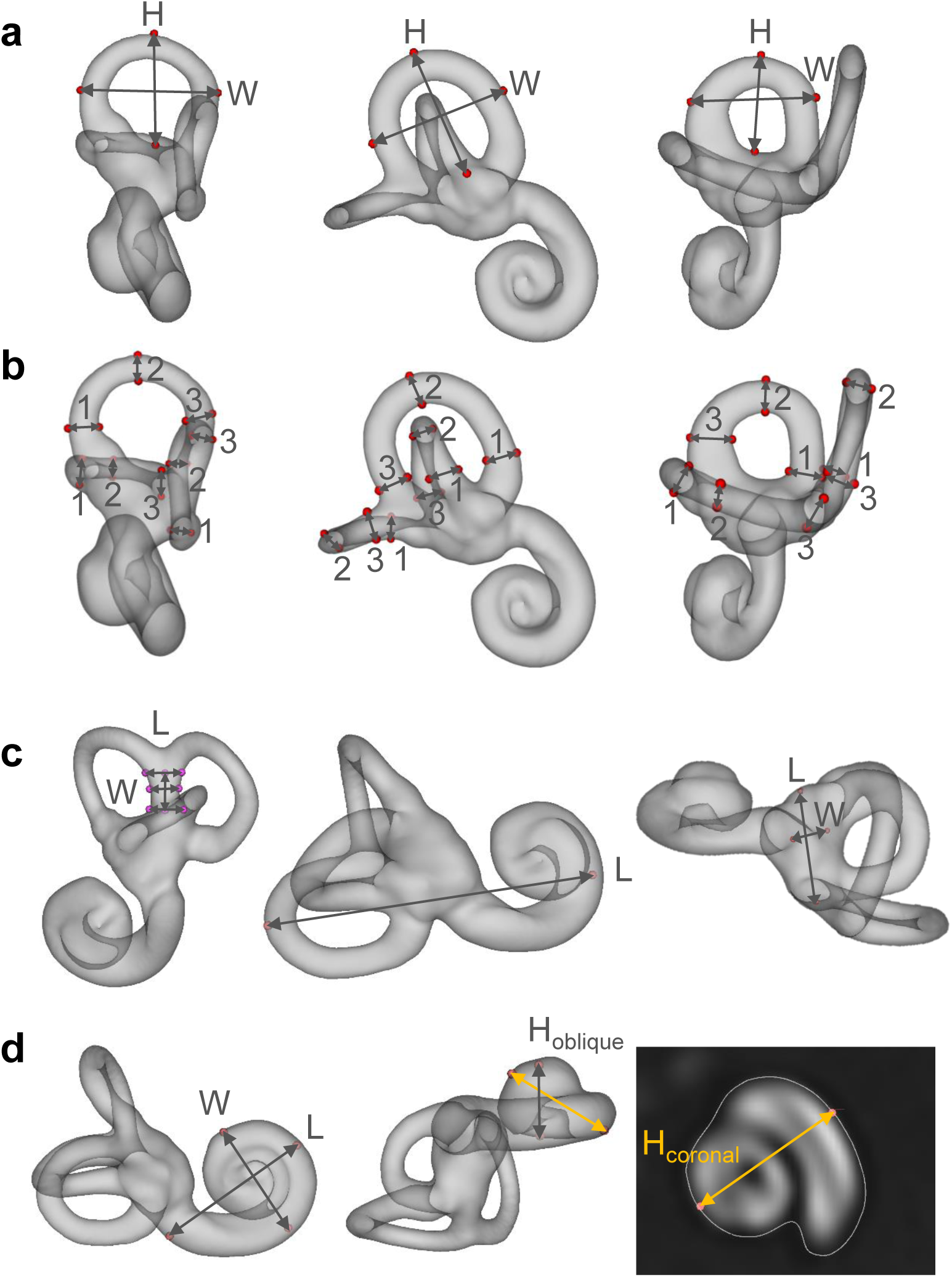
Inner ear landmark annotations in template space. Panel a: width and height of the anterior (left), posterior (middle) and lateral (right) semicircular canals. Panel b: inner widths and heights of semicircular canals, measured at three locations each (1-3). Panel c: height and three widths of the common crus (left), total inner ear length (middle), and vestibule length and width (right). Panel d: width, length and height of the cochlea. Cochlea height was tagged and measured using both the coronal plane approach (gold arrow)^51,52^, and the oblique plane (i.e. 3D) approach (grey arrow)^53^. H = height, W = width, L = length.

### Statistical methods

First-order group statistics on inner ear dimensions and volumes were computed in terms of mean and standard deviation (SD). Normality of variables throughout this work was tested using a Kolmogorov-Smirnov (KS) test with Lilliefors correction^72^, at an alpha level of *p <* 0.05. Comparisons of the semicircular canal dimensions between the left and right inner ear sides were performed using multiple paired statistical tests (two-tailed t-test for normal distributed data, Mann-Whitney U test for non-normal distributions). We corrected p-values after multiple testing using the Benjamini-Hochberg False Discovery Rate (FDR) algorithm^73^, results were considered as significant at a post-correction alpha level of *p* < 0.05. For the main general inner ear measures (total inner ear volume, total inner ear length, oblique cochlea height, and radii of anterior/posterior/lateral SCCs), we investigate a potential correlation with overall head size, represented by TIV. Correlation is reported as Pearson’s correlation coefficient *r*, again at a significance level of *p* < 0.05. After clarification of the role of TIV, we tested gender effects, i.e. whether there was a significant difference between male and female cohort subgroups. These tests were performed on the two major inner ear measures, total inner ear volume and total inner ear length. To account for head size, we performed a One-way Analysis of Covariance (ANCOVA), controlling for TIV as the relevant covariate^74^. Statistical computation was performed using python implementations in scipy.stats^75^, Pingouin^76^ and Statsmodels^77^, as well as MATLAB (The Mathworks Inc., MA, USA). Where available and applicable, we provide comparable values from literature that we are aware of, by referencing reported mean and standard deviation values.

## Data availability

The atlas material is publicly available for download in our project repository on github. To download, follow instructions at: https://github.com/pydsgz/IEMap.

## Acknowledgements

The study was supported by the German Federal Ministry of Education and Research (BMBF) in connection with the foundation of the German Center for Vertigo and Balance Disorders (DSGZ) (grant number 01 EO 0901), and a stipend of the Graduate School of Systemic Neurosciences (DFG-GSC 82/3) to Theresa Marie Raiser. We thank Paul Taylor for critical input and feedback.

## Author contributions statement

PzE designed the study together with SSA and TMR. TMR and RMR collected the data under the supervision of VLF and PzE. Data analyses were done by SAA. SAA and TMR wrote the initial paper as shared first authors, all listed authors revised and acknowledged the final version.

## Additional information

### Competing interests

The authors declare no competing interests.

## Notes

### Competing Interest Statement

The authors have declared no competing interest.

